# Use of tobacco and nicotine products among adolescents in Sub-Saharan Africa: protocol for a population-based multi-country household survey

**DOI:** 10.1101/2024.05.21.24307670

**Authors:** Lyagamula Kisia, Shukri F Mohamed, Grace Kyule, Christelle Tchoupé, Olatunbosun Abolarin, Retselisitsoe Pokothoane, Terefe Gelibo Agerfa, Samuel Iddi, Boscow Okumu, Nelson Mbaya, Damazo T Kadengye, Noreen Dadirai Mdege, Data on Youth and Tobacco in Africa (DaYTA) consortium

## Abstract

**Background:** The use of tobacco among adolescents in low- and middle-income countries, is a public health issue of concern. The tobacco industry’s aggressive marketing tactics target young people in African countries, leading to early initiation of tobacco use. While existing evidence focuses on 13-15-year-olds, data from Sub-Saharan Africa indicates that smoking initiation ranges from as young as 7 years old to around 16 years old. The lack of data on adolescent tobacco use in African countries limits policymakers’ ability to implement evidence-based tobacco control policies. This study aims to address the critical lack of quality and timely primary data on adolescent tobacco use thereby enhancing the country’s capacity to target interventions effectively, engage local governments, and attract global attention and funding for adolescent health initiatives.

**Methods:** We will conduct a cross-sectional nationwide survey among adolescents aged 10 - 17 years in urban and rural areas of the Democratic Republic of Congo (DRC), Kenya and Nigeria. This household-based survey will utilise a multi-stage stratified sample design to ensure representation across diverse geographic and demographic characteristics.

**Discussion:** Through this initiative, we aim to catalyse action at national and international levels to combat the tobacco epidemic among adolescents in SSA. The findings from the DaYTA study will empower stakeholders to advocate for effective tobacco control measures, promote adolescent health, and safeguard future generations from the harmful effects of tobacco use.

## Background

The Data on Youth and Tobacco in Africa (DaYTA) program is designed to address critical data gaps in tobacco control, focusing on tobacco use among adolescents in Sub-Saharan Africa (SSA). This region has one of the youngest populations globally, for instance, 57.6% of the population in the Democratic Republic of Congo (DRC) and 52% of the population in Nigeria is under 19 years of age [1, 2]. The tobacco industry targets these young populations through aggressive marketing strategies, including celebrity endorsements, advertising near schools and playgrounds, distributing free products, and producing youth-oriented flavours such as those in tobacco products like shisha (hookah/waterpipe) which were traditionally uncommon among young people but are increasingly becoming popular in this population. Additionally, the production of ‘Novel’ tobacco and nicotine products such as e-cigarettes present another opportunity for risky behaviour. These trends are attributed to the perception among many adolescents that these products are ‘safer’ than cigarettes [3– 5]. Evidence suggests that in addition to the immediate health risks associated with the use of these products, adolescents who use e-cigarettes/vape are three times more likely to have ever smoked combustible cigarettes and twice more likely to be current smokers [6, 7]. Those who start using e-cigarettes earlier in their adolescence are also more likely to use cigarettes later in life than those who start using them later [8]. Yet, data on use of these products in Africa remains sparse and policy interventions are lagging behind and are often based on regulation of traditional tobacco products.

Most data on adolescent smoking in SSA are derived from school-based surveys such as the Global Youth Tobacco Survey (GYTS) and Global School-based Student Health Surveys (GSHS), which target 13–15-year-olds. However, these studies have notable limitations: they exclude out-of-school adolescents and do not capture early (i.e., ages 10 years) and late (i.e., ages 18-19 years) adolescents [9]. Evidence suggests that smoking initiation can occur as early as 7 years old in some SSA countries [8, 10]. In SSA, a substantial proportion of children are not enrolled in school, with 20% of those aged 6-11 years, 33% of 12-14 years, and 48% of 15-17 years currently out of school [9]. Research indicates that out-of-school youth are more likely to initiate smoking than those who are in school [11, 12]. Furthermore, there are concerns about social desirability bias in school-based surveys where tobacco use is prohibited. In some SSA countries such as Kenya, tobacco is the most widely known and used psychoactive substance among those aged 14 years and younger in primary school [13].

Additionally, school-based data in many SSA countries is outdated, with the latest GYTS in Nigeria and DRC conducted in 2008, covering limited areas and missing a significant proportion of adolescents [14, 15]. In Kenya, the most recent nationally representative survey on tobacco use among 11-17-year-olds is more than a decade old [16], hindering evidence-based policy development and tobacco control interventions.

The DaYTA program seeks to bridge these gaps by conducting a population-based household survey in the DRC, Kenya, and Nigeria. This survey will capture data on tobacco and nicotine product use among adolescents aged 10-17 years, ensuring a comprehensive and timely assessment of tobacco trends. By focusing on household surveys, this protocol aims to include out-of-school adolescents and provide nuanced data that can inform evidence-based policy and intervention strategies for tobacco control in SSA.

## Methods/Design

### Aim

The primary goal of this study is to collect bespoke, nationally representative data in the DRC, Kenya, and Nigeria on tobacco and nicotine product use among adolescents aged 10 - 17 years, aiming to fill critical evidence gaps and complement existing data. We will address the following specific research questions:

1. What is the prevalence of tobacco and nicotine product use among adolescents aged 10-17 years?
2. What are the multi-level (e.g., individual-, household- and environment-level) factors associated with tobacco and nicotine product use among adolescents?

### Design and study population

A nationally representative population-based household survey will be conducted in each of the three countries. The surveys will focus on adolescents aged 10 - 17 years and their caretakers.

#### Eligibility criteria

To be eligible for inclusion, households must have at least one adolescent aged 10 - 17 years. In this study, a household is defined as a person or group of related or unrelated persons who live together in the same dwelling unit(s), who acknowledge one adult male or female as the head of the household, who pool some, or all, of their income and wealth, who consume certain types of goods and services collectively, mainly housing and food and who are considered a single unit [17, 18]. Household participation requires consent from the head of the household.

An adolescent will be eligible for the study if they are aged 10 - 17 years and are a member of a participating household. We will exclude adolescents who are unable to consent due to either refusal or inability to comprehend study information, adolescents who do not have the capacity to understand the questions being asked, and those with significant physical disabilities (e.g. hearing and speech impairment) that prevent the interviewer from oral administration of the surveys.

### Study Settings

The survey will be conducted in three SSA countries (DRC, Kenya, and Nigeria). The DRC is the largest country by land size in SSA. The study will be implemented in 16 randomly sampled from its 26 current provinces. According to DRC’s health pyramid, each province is divided into health zones (well-defined geographical areas, within the territorial boundaries of a municipality or territory, with a population of approximately 50,000 to 100,000 inhabitants in rural areas and 150,000 to 200,000 inhabitants in urban areas), and each health zone into health areas (geographical area with a population of approximately 5,000 inhabitants in rural areas and 10,000 inhabitants in urban areas) [19]. Kenya is a devolved system of governance made up of 47 county governments. The administrative structure of the counties comprises sub-counties, wards, and villages. In Kenya, the study will be implemented in 16 counties, that is, 15 randomly sampled counties and Nairobi which has been selected with certainty as it is the capital and largest city. On the other hand, Nigeria is divided into six geopolitical zones based on the 36 states. Each state is divided into Local Government Areas (LGAs), and the LGAs further divided, into localities. The study will be implemented in 13 states, where 12 will be randomly selected and the Federal Capital Territory (FCT) will be selected with certainty due to its cultural and ethnic diversity. It also hosts Abuja which is Nigeria’s administrative and political capital.

### Sampling Procedures

In each country, the survey will utilise a multi-stage stratified cluster sample design, with slight modifications taking into consideration the context of each country. In Kenya and Nigeria, stratification will be carried out based on the administrative divisions and the sampling frames will be obtained from the Kenya National Bureau of Statistics (KNBS) and the National Population Commission (NPC) in Kenya and Nigeria respectively. The lists will consist of data that allow multi-stage sampling, for example, information on administrative structures (e.g., Zones/States/Regions/Counties/Provinces, residential areas (Rural/Urban), and enumeration areas (EA)) and the relevant bodies will also provide maps and where available the list of households in each EA (cluster) in the selected counties. In the absence of household listing, household listing will be carried out to identify eligible households. In DRC, stratification will be carried out based on the health pyramid structure delineated by the Ministry of Public Health, Hygiene and Prevention into health zones, and health areas. Avenues/villages within the health area will serve as clusters. The sampling frame will be obtained from the National Program for Countering Drug Addiction and Toxic Substances (PNLTC). The health pyramid will serve as the sampling framework and the list and maps covering the country will be obtained from the National Institute of Statistics (INS). Household listing will be carried out to identify eligible hosueholds.

#### Sampling strategy for DRC

In the DRC, the first stage will involve stratified random sampling of the provinces. Stratification will be based on the six former provinces of DRC between 1947 to 1963 which include Katanga, Kasaï, Léopoldville, Équateur, Orientale, and Kivu. In the second stage, three Health Zones (HZs) (one in an urban area and two in rural areas) will be randomly selected in each of these provinces. In the third stage, three Health Areas (HAs) will be selected in each of the participating HZs. For the fourth stage, in the HAs, one avenue or village will be selected randomly according if the HA is in the urban areas, or rural areas. In the fifth stage, in each of these avenues/villages, a proportion of households (and therefore of one adolescent aged 10 to 17 per household) will be randomly selected and surveyed. This systematic approach will guarantee a comprehensive and representative coverage of different geographic strata within the DRC. For consistency across the countries, it is important to note that avenue or village in DRC will be referred to as EAs like in Kenya and Nigeria.

#### Sampling Strategy for Kenya

In Kenya, the first sampling stage will involve selecting counties from the county sampling frame. The second stage will involve random sampling of EAs from the selected counties with a probability proportional to the size of the sampled counties. The survey team will carry out a household listing operation in all selected EAs before the start of fieldwork. The household list will serve as the sampling frame for the third stage of sample selection, where a fixed number of households (30) to be interviewed will be selected from each EA using a systematic random sampling technique. In each of the selected households, one adolescent aged 10 to 17 will be interviewed.

#### Sampling strategy for Nigeria

The sampling strategy in Nigeria is like that of Kenya. The first stage involves selecting states from the state sampling frame. The second stage involves randomly of EAs from the selected states and finally selecting a fixed number of households (30) from the selected EAs in each state.

### Participant recruitment

The survey team will conduct a household listing within the selected EAs working closely with the local and community leaders/representatives to demarcate EA boundaries. Within these boundaries each household will be visited to identify if there is an adolescent aged 10-17 years living in the house who is available to participate in the survey. This process will establish a sampling frame from which eligible households will be randomly selected for inclusion. The survey teams will obtain consent from the head or acting head of the randomly selected household prior to administering the household survey. During the household survey, a household roster will be populated, and all eligible adolescents within the household will be identified. In households with more than one eligible adolescent, one will be randomly selected for participation in the study. Parental consent and adolescent assent will be sought prior to administering the adolescent survey questionnaire. For emancipated minors (those living independently from parents and competent to make their own decisions), consent will be sought directly from the adolescent prior to the questionnaires being administered.

### Sample size calculations

Sample size calculations will adhere to established methodologies tailored to each country. The United Nations (UN)’s formula for prevalence studies [20] will guide sample size computation in Kenya and Nigeria, while the DRC’s formula will be adapted from the Multiple Indicator Cluster Survey (MICS)-Palu RDC 2017–2018 [1], with a confidence level of 95% for all the countries. The sample design effect will be set at 2 for Kenya, 2.5 for Nigeria, and 1.5 for the DRC, and considering non-response rate of 10% for Kenya and the DRC [21], and 20% for Nigeria. The estimated level of tobacco prevalence for the countries are based on each country’s most recent estimates from recent studies. The adolescent population proportions are estimated based on national statistics from both countries and are 20.45% for Kenya [22], 17.9% for Nigeria [23], and 23% for the DRC [15]. The average household size is 3.9[22], 4.7 [23], and 5.25 [1] for Kenya, Nigeria, and the DRC, respectively. This will result in a nationally representative samples of 6,701 adolescents in Kenya, 4,803 adolescents in the DRC, and 7,948 adolescents in Nigeria.

### Questionnaire development process

The DaYTA standardised questionnaire was developed through intensive review of literature, drawing insights from internationally recognized survey tools such as the CDC National Youth Tobacco Survey (NYTS) [24]; The GYTS [25]; Global Adult Tobacco Survey (GATS) [26]; ASH Smoke free Great Britain Youth Survey (ASH-Y) [27]; International Tobacco Control (ITC)-Youth Surveys [27]; WHO Tobacco Questions for Surveys of Youth (TQS-Youth) [28]. This review process was complemented by consultations with key country stakeholders (Figure 2), involving one-on-one interviews to understand their data needs and priorities for decision-making. Stakeholder input guided the questionnaire drafting process, culminating in country-level workshops where the questionnaire was presented and refined based on feedback from local stakeholders. A cross-country workshop further ensured the relevance and appropriateness across the three participating countries. The questionnaires underwent rigorous field testing before finalization and use.

**Figure 2:**
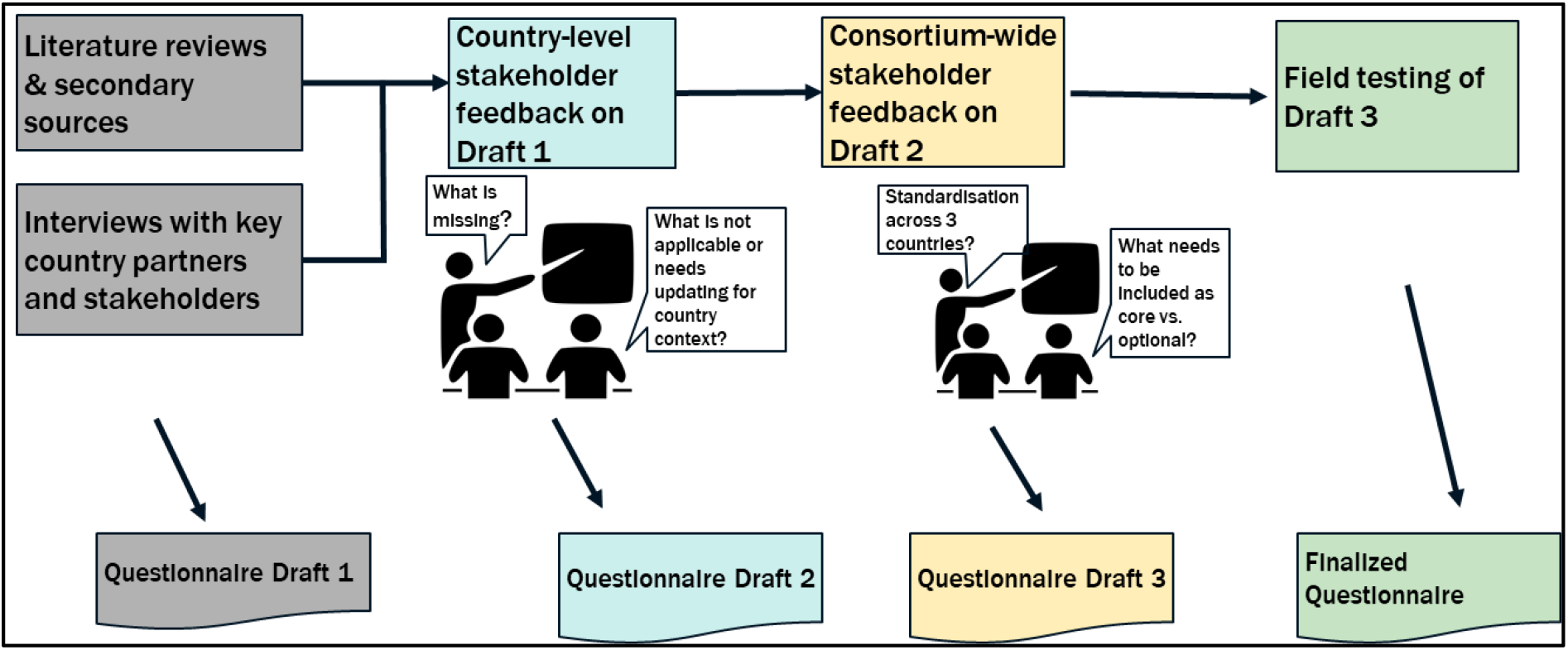
Questionnaire development process.

### The household questionnaire

will be administered to the consenting head of household or acting head of household and consists of two distinct modules focusing on demographics and socio-economic status. The first module, a household roster, will collect demographic details including sex, age, income, disability status, marital status, health insurance cover and education of the de facto members of the household. The second module will collect information on household characteristics pertinent to socio-economic assessment such as sources of drinking water, access to sanitation and cooking facilities, housing structure and materials, and ownership of assets.

### The adolescent questionnaire

will be administered to participating adolescents and includes 12 modules aimed at collecting socio-demographic characteristics, tobacco and nicotine product use behaviours, and multi-level (e.g., individual-, household- and environment-level) factors associated with tobacco use and nicotine products. The questionnaire will cover the following:

- Socio-demographic characteristics such as age, sex, school year (if in school), average weekly spending money, in-school/ out-of-school, parents/guardians/other family members’ tobacco use histories, and tobacco use amongst close friends; functional difficulties i.e. vision, mobility, cognition remembering, self-care and communication.
- Use of smoked tobacco (manufactured/factory-made cigarettes, roll-your-own (RYO)/hand-rolled cigarettes, shisha/waterpipe/hookah, and other smoked tobacco products e.g. cigars, cheroots, cigarillos), heated tobacco products, smokeless tobacco (chewing tobacco such as tobacco leaf, tobacco leaf, and lime; kuber, applying tobacco such as tobacco toothpaste-dentobac etc.; tobacco tooth powder-lal, etc.; snuff), electronic cigarettes, and nicotine pouches. For each product, or product type, we will collect information including quantity, frequency, dependency, age of initiation, where they smoke, and with whom, and access (how they access, where and for how much)
- Knowledge, attitudes, perceptions, and intentions regarding tobacco use and its consequences including exposure to tobacco advertising, promotion or sponsorship, and exposure to anti-tobacco messages.
- Information on cessation of tobacco use (for those using tobacco products), and second-hand exposure to tobacco smoke within the home and in indoor and outdoor public places.

### Translations and back translations of study documents

Translations and back-translations of survey tools and consent/assent forms will be conducted to ensure linguistic accuracy and to maintain the integrity of the content. First, the survey tools and consent/assent forms will be translated from English into the identified language(s) by a professional translator who is a native speaker. Subsequently, a different native speaker of the language(s) will perform a back translation, without prior knowledge of the original text, to verify that the intended meanings were preserved and accurately presented. Instruments will be harmonised to ensure consistency and coherence in the textual content across all survey materials.

### Data collection process

Data will be collected by trained field interviewers with prior experience working with similar surveys and who possess knowledge of the local context and selected languages in each country. Experienced researchers will supervise the data collection process. Data collection will be done in person using an interviewer-administered electronic questionnaires programmed in Survey CTO on tablets. Data will be transmitted to online secure servers for storage after all quality checks are completed. Interviews with adolescents will be done in a private setting to avoid interference from parents or caregivers. Parents will be informed during the consenting process the topics that will be discussed with the adolescents and advised not to interfere once consent is given. Interviewers will be advised to end the interview if privacy cannot be maintained.

To ensure improved data quality, an electronic questionnaire will include appropriate skip-logic patterns will be programmed into the electronic data collection devices and spot checks will be conducted on at least 5% of the sample to verify data accuracy. Field interviewers will ensure that every question has been asked and that responses are recorded clearly and accurately before completing each interview. Regular data validation and verification checks will also be run on all the data collected using a syntax script to ensure data completeness, correctness, and consistency. Supervisors will maintain regular communication with the central coordination team to discuss progress and address any operational challenges, facilitating adjustments to the data collection process as needed.

### Data management

The data will be collected using the offline module and will be uploaded onto a secure server using internet connectivity or mobile data. Backup of the data will remain on the tablets until the end of field activities. Data transmitted to the central servers will be password protected to allow access to only authorized users. Households and individuals who have consented to participate in the study will be assigned a unique study identification number. This identification number will be associated with all participant’s data that is collected, entered, and analysed for the study. To ensure confidentiality, all personal identifiers (name, identity numbers, phone numbers and places of residence) collected during data collection or for recruitment procedures will be removed from analytical datasets before any data is shared or used in analysis. The raw data will be cleaned and transformed as needed for the statistical analysis. The codes for this purpose will be written in do-files from STATA or R-scripts, to allow traceability and verification of the cleaning operations carried out. This process will involve identifying and addressing missing values, outliers, responses coded as ‘other’ and any data inconsistencies. With the clean data, we shall produce detailed reports with completed tables on different variables as well as a more condensed summary of the results.

### Statistical analysis

Descriptive statistics will be used to explore the prevalence and distribution of tobacco use among adolescents at the country and county/province/state levels, including disaggregation by variables of interest such as sex, in-school/out-of-school, rural/urban, and socioeconomic differences, and other household- or person-level characteristics. Survey weighted proportions and percentages, means, medians, and standard deviations of variables will also be computed and presented. Visualisation techniques such as graphs and charts will be used to represent the variables of interest and results of the analysis to help in communicating key findings and insights effectively.

Since the survey is a multi-stage stratified cluster sample, initial sampling weights will be calculated by multiplying the inverse of the probability of selection at each stage of the sampling plan.

Adjustments to the weight will be made to account for non-response and calibration adjustment factors and thus, final weights will be computed by multiplying the initial weights, the non-response adjustment factor, and the calibration or post-stratification factor for each sampled unit. The final weights will be normalized to match population totals.

Survey weighted multivariable models (i.e., logistic regression) will be fitted to estimate adjusted odds ratios for the relationships between tobacco use with other explanatory variables such as age at initiation, smoking cessation attempts, the intensity of tobacco use(quantity consumed), frequency of use and type of the tobacco product; while controlling for individual factors (e.g., age, sex, ethnicity, education level, knowledge/perceptions about tobacco products), household factors (e.g., household size, family structure, wealth index) and environment-level factors (e.g., residence (rural/urban), geographical location (region/county/state), exposure to tobacco advertising, access to the products). These statistical techniques will account for the sampling design (stratification and clustering) and the computed sampling weights.

## Discussion

The study seeks to fill critical gaps in knowledge regarding tobacco use among adolescents aged 10-17 years in three SSA countries: the DRC, Kenya, and Nigeria.

The findings from the study have the potential to significantly influence both policy and public health initiatives related to adolescent tobacco use in the DRC, Kenya, and Nigeria. To enable this, the study findings will be communicated at multiple levels to ensure broad impact and engagements with the stakeholders in the tobacco control space across the DRC, Kenya, and Nigeria. We will engage with diverse stakeholders including Ministries of Health, program managers, community and youth representatives, advocacy groups, civil society groups, and non-governmental organizations. At the end of the study, a stakeholders meeting will be organized to share the findings with the DaYTA teams and stakeholders. Country-level dissemination meetings will also be conducted to engage stakeholders in strategy sessions, discuss actionable next steps, and disseminate key findings from the study report. Additionally, the study findings will be disseminated through peer-reviewed journal publications, and scientific conferences and made accessible on the Tobacco Control Data Initiative (TCDI) dashboard, https://tobaccocontroldata.org.

### Strengths of the study

The three study countries (DRC, Kenya, and Nigeria) are diverse in terms of ethnic, linguistic, ecological, environmental, religious, and other contexts, representing the East, Central, and Western regions of Africa. This diversity offers a novel empirical contribution by examining a range of distinct but independent dimensions of field operations. These dimensions include not only data collection and capacity but also the challenges and experiences of conducting surveys, as well as vital questions of context and diversity. Future studies could benefit, explore, and compare alternative methodologies in terms of coverage, mode of data collection, frequency, geographical detail, response rate, quality, cost, required resources, and timeliness.

### Study limitations

While this study is designed to maximize, reach, and impact, there are inherent limitations to be acknowledged. First, despite the intention to conduct a nationwide survey, logistical challenges could impede, potentially leading to data representation access to certain remote and crisis-affected areas, potentially leading to data representation biases. Secondly, reliance on self-reported information introduces the risk of biases, as participants might underreport or over-report their tobacco use due to social desirability and recall errors.

### Feasibility and logistical considerations

Implementing a survey of this magnitude requires careful consideration of feasibility and logistical challenges. The success of such a survey hinges on meticulous planning and allocation of substantial resources. Securing the cooperation of local communities, and managing the nationwide survey, in diverse settings can pose significant challenges such as a lack of sufficient initial buy-in from key stakeholders. To mitigate this, we conducted country-specific assessments where we established that government officials and other key stakeholders were concerned about youth and the lack of adolescent data. We also conducted cross-country workshops to validate this concern and to gather more information. As is possible with all political engagements, shifting priorities and/or shifts in government positions may lead to what began as strong initial buy-in shifting to less buy-in and engagement. The program will continue to work closely with the government to ensure they continue to provide feedback at each stage of the research, provide national and local context for intervention activities, and guide dissemination activities. We hope that this will sustain high-level political buy-in. Cementing strong relationships with government ministries and institutions, and other key stakeholders is also critical to ensuring high levels of trust in the data created, curated, and shared through this program.

Another challenge is the insecurity in certain areas that may hinder data collection and pose a significant risk to the field teams and investigators. Such areas were identified in collaboration with local authorities and relevant bodies and removed prior to the selection of EAs. It may also be difficult to gain access to some of the selected EAs. This obstacle will be addressed through robust community mobilization strategies and the engagement of relevant local authorities to facilitate smooth operations and ensure the validity of the collected data.

### Further research directions

The study lays the groundwork for future longitudinal studies aimed at tracking changes in tobacco use patterns over time among adolescents. Subsequent research could delve into evaluating the effectiveness of targeted anti-tobacco interventions that are informed by the data collected through this survey. Additionally, qualitative studies could unpack life experiences or contextual factors that shape tobacco and nicotine product use behaviours. Replicating this study in other regions could provide a broader perspective on youth tobacco use, contributing to a more comprehensive foundation for global health interventions aimed at curbing tobacco use among young populations.

### Conclusion

This study protocol outlines a structured and strategic approach to investigating tobacco use among African adolescents. Despite the limitations, the strength of the study lies in its comprehensive designs and potential to impact public health policy significantly. By advancing this study, the researchers aim to equip stakeholders with the data necessary to develop effective, sustainable, and culturally sensitive anti-tobacco interventions tailored to youth populations across Africa.

## Data Availability

The datasets generated and/or analysed during the current study will be available on the TCDI
dashboard, https://tobaccocontroldata.org. The datasets generated and/or analysed for Kenya during
the current study will also be available on the APHRC microdata portal,
http://microdataportal.aphrc.org/

## List of abbreviations

ASH-Y: ASH Smoke free Great Britain Youth survey (ASH-Y)
CDC NYTS: Centers for Disease Control and Prevention National Youth Tobacco Survey
DaYTA: Data on Youth and Tobacco in Africa
DRC: Democratic Republic of Congo
EA: Enumeration Area
FCT: Federal Capital Territory
GATS: Global Adult Tobacco Survey
GSHS: Global School-based Student Health Survey
GYTS: Global Youth Tobacco Survey
INS: National Institute of Statistics
ITC-Youth: International Tobacco Control (ITC)-Youth Surveys
KNBS: Kenya National Bureau of Statistics
MICS: Multiple Indicator Cluster Survey
SSA: Sub Saharan Africa
NPC: National Population Commission
PNLTC: National Program for the Fight against Drug Addiction and Toxic Substances
RYO: Roll-your-own
TAPS: Tobacco Advertisement, Promotion, and Sponsorship
TCDI: Tobacco Control Data Initiative
UN: United Nations
WHO: TQS World Health Organisation Tobacco Questions for Surveys of Youth

## Declarations

### Ethics approval and consent to participate

Each country obtained ethical approval from the relevant national ethical bodies. In DRC, ethical approval (001/DIR/RISD/2024) was received from the National Health Ethics Committee. In Kenya, ethical approval (P1570/2023) was received from AMREF Health Africa’s Research Ethics and Scientific Review Committee (ESRC) and a research license (NACOSTI/P/24/32385) from the National Commission for Science, Technology and Innovation (NACOSTI). In Nigeria, ethical approval (01/01/2007 – 19/01/2024) was received from the National Health Research Ethics Committee of Nigeria (MNHREC). Each country sought out additional approvals where necessary.

### Consent for publication

Not Applicable

## Availability of data and materials

The datasets generated and/or analysed during the current study will be available on the TCDI dashboard, https://tobaccocontroldata.org. The datasets generated and/or analysed for Kenya during the current study will also be available on the APHRC microdata portal, http://microdataportal.aphrc.org/

## Competing interests

The authors declare that they have no competing interests.

## Funding

This work is supported by the Bill & Melinda Gates Foundation (INV-048743). The Bill & Melinda Gates Foundation had no role in the design of the study; in the collection, analyses, or interpretation of data; in the writing of the manuscript, or in the decision to publish the results. The findings and conclusions contained within are those of the authors and do not necessarily reflect positions or policies of the Bill & Melinda Gates Foundation.

## Authors’ contributions

NDM conceptualised and led the funding acquisition. LK drafted and led the writing of the manuscript. GK and SI contributed to revising the statistical aspects of the manuscript. OA contributed to writing the discussion section. NDM, SFM, DTK, CT, RP, TGA, BO provided critical feedback on various sections of the manuscript. NDM and SFM provided overall guidance on the content to all versions of the manuscript. All authors contributed to reading, revising, and approving the final manuscript.

## Acknowledgments

This study was conducted as part of the Data on Youth and Tobacco in Africa (DaYTA) program. The DaYTA program seeks to empower stakeholders to make timely, data-driven decisions by using evidence to inform policy and advance tobacco control efforts in sub-Saharan Africa. The program achieves this by addressing data gaps related to tobacco use among 10-17-year-olds in Kenya, Nigeria, and the Democratic Republic of Congo. The DaYTA program is led by Development Gateway: an IREX Venture (DG), a global non-profit organization that specializes in data for development.

We are grateful to the participants in each country for the time they took to participate in the study. We acknowledge the invaluable inputs from the DaYTA Program consortium members as follows:

## DRC

Patrice Milambo, National Program to Combat Drug Addiction and Toxic Substances (PNLCT); Dr. Célestin Banza Lubaba Nkulu, University of Lubumbashi; Dr. Josaphat Ndelo-di-Phanzu, University of Kinshasa; Hyppolite Phanzu Lusala, Local Initiative for Integrated Development (ILDI); Didier Munguakonkwa Mirindi, RISD; Emmanuel Kandate, RISD; Dr. Patrick Shamba, Development Gateway (DG); Christus Miderho (DG)

## Kenya

Dorcas Kiptui, Ministry of Health; Anne Kendagor, Ministry of Health; Dr. Lydia Mucheru, Kenya Institute of Curriculum Development (KICD); Elias Nyaga, Kenya National Bureau of Statistics (KNBS); Edwin Metto, Kenya National Bureau of Statistics (KNBS); Jimi Kirimi, Kenya National Bureau of Statistics (KNBS); Jackline Chepkorir, Kenya National Bureau of Statistics (KNBS); Fabian Oriri, International Institute for Legislative Affairs (IILA); Winnie Awuor, DG; Rachel Kitonyo-Devotsu, DG

## Nigeria

Dr. Malau Mangai Toma (head), Federal Ministry of Health NCD Division; Emmanuel Abraham, Federal Ministry of Health NCD Division; Emmanuel Davies, Federal Ministry of Health NCD Division; Dr. Adedeji Adeniran, Center for the Study of Economies of Africa (CSEA); Esther Aghotor, Gatefield; Philip Jakpor, Renevlyn Development Initiative (RDI); Chibuike Nwokorie, Nigeria Tobacco Control Alliance (NTCA); Uche Michael, APIN Public Health Initiatives, Akinsewa Akiode, RCS; Thompson Ademola, RCS; Mohammed Maikudi, DG

## Additional members

Lauren Eby, DG, Franklin Koech, APHRC; James Amukohe Kavai, APHRC

## Notes

### Competing Interest Statement

The authors have declared no competing interest.

### Author Declarations

Ethics committee/IRB of National Health Ethics Committee gave ethical approval for this work in the DRC. Ethics committee/IRB of AMREF Health Africa gave ethical approval for this work in Kenya. Ethics committee/IRB of National Health Research Ethics Committee gave ethical approval for this work in Nigeria.

## References

[1] INS, “Enquête par grappes à indicateurs multiples 2017 - 2018: rapport de résultats de l’enquête,” Kinshasa, DRC, 2019.

[2] D. Adeloye et al., “Current prevalence pattern of tobacco smoking in Nigeria: A systematic review and meta-analysis,” BMC Public Health, vol. 19, no. 1, pp. 1–14, Dec. 2019, doi: 10.1186/S12889-019-8010-8/FIGURES/4.

[3] A. E. O. Ogwell, A. N. Aström, and O. Haugejorden, “Socio-demographic factors of pupils who use tobacco in randomly-selected primary schools in Nairobi province, Kenya,” East Afr. Med. J., vol. 80, no. 5, pp. 235–241, May 2003, doi: 10.4314/EAMJ.V80I5.8693.

[4] W. K. Maina, J. N. Nato, M. A. Okoth, D. J. Kiptui, and A. O. Ogwell, “Prevalence of Tobacco Use and Associated Behaviours and Exposures among the Youth in Kenya: Report of the Global Youth Tobacco Survey in 2007,” Public Heal. Res., vol. 3, no. 3, pp. 43–48, Jan. 2013, doi: 10.5923/J.PHR.20130303.03.

[5] A. Olawole-Isaac, O. Ogundipe, E. O. Amoo, and D. O. Adeloye, “Substance use among adolescents in sub-Saharan Africa: a systematic review and meta-analysis,” South African J. Child Heal., vol. 12, no. 2b, p. 79, Sep. 2018, doi: 10.7196/SAJCH.2018.V12I2B.1524.

[6] P. B. James, A. J. Bah, J. A. Kabba, S. A. Kassim, and P. A. Dalinjong, “Prevalence and correlates of current tobacco use and non-user susceptibility to using tobacco products among school-going adolescents in 22 African countries: a secondary analysis of the 2013-2018 global youth tobacco surveys,” Arch. Public Heal., vol. 80, no. 1, pp. 1–15, Dec. 2022, doi: 10.1186/S13690-022-00881-8/TABLES/5.

[7] B. K. Oyewole, V. J. Animasahun, and H. J. Chapman, “Tobacco use in Nigerian youth: A systematic review,” PLoS One, vol. 13, no. 5, p. e0196362, May 2018, doi: 10.1371/JOURNAL.PONE.0196362.

[8] O. G. Chido-Amajuoyi, P. Fueta, and D. Mantey, “Age at Smoking Initiation and Prevalence of Cigarette Use Among Youths in Sub-Saharan Africa, 2014-2017,” JAMA Netw. Open, vol. 4, no. 5, May 2021, doi: 10.1001/JAMANETWORKOPEN.2021.8060.

[9] R. Desai, L. A. G. Mercken, R. A. C. Ruiter, J. Schepers, and P. S. Reddy, “Cigarette smoking and reasons for leaving school among school dropouts in South Africa,” BMC Public Health, vol. 19, no. 1, pp. 1–10, Jan. 2019, doi: 10.1186/S12889-019-6454-5/TABLES/4.

[10] S. P. Veeranki et al., “Age of smoking initiation among adolescents in Africa,” Int. J. Public Health, vol. 62, no. 1, pp. 63–72, Jan. 2017, doi: 10.1007/S00038-016-0888-7/METRICS.

[11] C. W. Warren, N. R. Jones, M. P. Eriksen, and S. Asma, “Patterns of global tobacco use in young people and implications for future chronic disease burden in adults,” Lancet (London, England), vol. 367, no. 9512, pp. 749–753, Mar. 2006, doi: 10.1016/S0140-6736(06)68192-0.

[12] World Health Organization, “Global Youth Tobacco Survey: Fact Sheet Mauritius 2016,” 2017.

[13] M. Kamenderi, J. Mutetei, V. Okioma, S. Kimani, F. Kanana, and C. Kahiu, “Status of Drugs and Substance Abuse in Kenya,” African J. Alcohol Drug Abus., vol. 1, pp. 54–59, 2019, Accessed: Nov. 16, 2023. [Online]. Available: https://www.researchgate.net/publication/345805433_Status_of_Drugs_and_Substance_Abusein_Kenya

[14] GYTS Lagos, “Global Youth Tobacco Survey (GYTS) for Nigeria,” 2008. Accessed: Nov. 16, 2023. [Online]. Available: https://extranet.who.int/ncdsmicrodata/index.php/catalog/793/related-materials

[15] GYTS DRC, “Global Youth Tobacco Survey (GYTS) for DRC,” 2008.

[16] GYTS Kenya, “GYTS Kenya: Fact Sheet 2013,” 2013.

[17] “UNdata | glossary,” UN. Accessed: Nov. 16, 2023. [Online]. Available: http://data.un.org/Glossary.aspx?q=household

[18] T. N. Croft, A. M. J. Marshall, and C. K. Allen, “Guide to DHS Statistics,” Rockville, Maryland, USA, 2018.

[19] C. Kayembe and M. Gyenano, “RD Congo: Etude auprès des infrastructures sanitaires des facteurs à la base de la mortalité à Kinshasa - Democratic Republic of the Congo | ReliefWeb,” OCHA. Accessed: May 20, 2024. [Online]. Available: https://reliefweb.int/report/democratic-republic-congo/rd-congo-etude-auprès-des-infrastructures-sanitaires-des-facteurs-à

[20] United Nations, Studies in Methods: Designing Household Survey Samples: Practical Guidelines, 98th ed., vol. F. New York, 2008.

[21] Ecole de Santé Publique de Kinshasa, “ENQUETE DE COUVERTURE VACCINALE CHEZ LES ENFANTS DE 6-23 MOIS En République Démocratique du Congo 2021,” 2021.

[22] Kenya National Bureau of Statistics, “2019 KENYA POPULATION AND HOUSING CENSUS: Volume 1,” 2019.

[23] National Population Comission (NPC), “2018 Demographic and Health Survey Key Findings Nigeria,” 2019. Accessed: May 13, 2024. [Online]. Available: https://www.population.gov.ng

[24] C. for Disease Control, “National Youth Tobacco Survey (NYTS) 2022 Questionnaire Thank You Very Much For Your Help,” 2022.

[25] Global Youth Tobacco Survey Collaborative Group, “Global Youth Tobacco Survey (GYTS) Core Questionnaire with Optional Questions GYTS Core Questionnaire with Optional Questions GYTS Sample Design and Weights GYTS Implementation Instructions GYTS Analysis and Reporting Package GYTS Data Dissemination Guida,” Atlanta, GA: U.S, 2023.

[26] Global Adult Tobacco Survey Collaborative Group, “Global Adult Tobacco Survey (GATS) Core Questionnaire with Optional Questions,” Atlanta, GA: U.S, 2020.

[27] Action on Smoking and Health (ASH), “Use of e-cigarettes (vapes) among young people in Great Britain,” 2023.

[28] World Health Organization and U.S. Centers for Disease Control and Prevention, “Tobacco Questions for Surveys of Youth (TQS-Youth) A Subset of Key Questions from the Global Youth Tobacco Survey (GYTS),” Geneva, 2019.

